# Gender-Specific Osteoporosis Risk Prediction Using Longitudinal Clinical Data and Machine Learning

**DOI:** 10.64898/2026.02.13.26346244

**Authors:** Shyama P. Tripathy, Lohitha Saripalli, Katherine Berry, Ambalangodage C. Jayasuriya, Devinder Kaur, Fayeq J. Syed

## Abstract

Osteoporosis is a silent yet debilitating disease that often remains undetected until fractures occur. While early prediction is crucial, most studies combine male and female datasets to train a single model, introducing bias since osteoporosis risk and progression differ by gender. This study aims to develop gender-specific machine learning models that leverage longitudinal data to predict osteoporosis risk, providing tailored insights for men and women. Data were obtained from two large longitudinal cohorts: the Study of Osteoporotic Fractures (SOF) for women and the Osteoporotic Fractures in Men Study (MrOS) for men. Multiple ML algorithms were trained and evaluated for each sex, with model performance assessed using the area under the receiver operating characteristic curve (AUC-ROC). Among the tested models, the XGBoost model demonstrated the best performance for women, achieving an AUC-ROC of 0.93 using SOF data. For men, the Random Forest model achieved an AUC-ROC of 0.89 using MrOS data. Feature importance analysis identified sex-specific osteoporosis risk factors, underscoring the need for tailored prediction and management. By revealing male and female risk factors and reducing bias from combined datasets, the work advances personalized care and supports earlier, effective clinical intervention to prevent fractures and improve health outcomes.

## 1. Introduction

Osteoporosis is defined by reduced bone amount and decreased bone mineral density (BMD), leading to a higher risk of fragility fractures that significantly impact quality of life and increase morbidity, mortality, and disability [1]. Affecting millions worldwide particularly in industrialized regions such as the United States, Europe, and Japan osteoporosis accounts for more than 8.9 million fractures annually, with a lifetime fracture risk of 30% in developed countries, comparable to that of coronary heart disease [2]. These fractures place substantial strain on individuals, families, and healthcare systems, and with aging populations and rising treatment costs, effective prevention and management strategies are increasingly essential to reduce long-term consequences and improve patient outcomes [3].

Although osteoporosis is often thought to primarily affect older women, it also impacts men and younger individuals, and it is often undiagnosed until a fracture leads to medical attention. Thankfully, osteoporosis can be treated and prevented, but only if detected early [4]. In order to manage the illness and enhance results, early detection is crucial. A comprehensive medical history, physical examination, specialized tests and timely intervention can help prevent further bone loss, reduce fracture risk, and improve the quality of life for those with osteoporosis [5].

Numerous studies have identified shared risk factors for osteoporosis, such as age, being female, a family history of the condition, low body mass index, various physical function factors, and lifestyle factors such as smoking and excessive alcohol consumption [6].This study makes use of longitudinal data from two significant datasets; the Study of Osteoporotic Fractures (SOF) [7] and the Osteoporotic Fractures in Men Study (MrOS) [8] to track the progression of osteoporosis over time. Longitudinal data, collected over multiple visits spanning several years, provides important insights into how key predictors including clinical data such as age, weight, fall history, physical functions, and diagnostic data involving BMD and bone mineral content (BMC), provide a deeper understanding of the progression of osteoporosis and the factors influencing fracture risk.

Machine Learning (ML) offers several advantages over traditional statistical methods, particularly for longitudinal data where assumptions may not hold and missing values are common [9]. ML models can capture complex temporal patterns and nonlinear relationships, making them effective for progressive diseases such as osteoporosis. Ensuring generalizability across populations is essential, as demonstrated in applications such as predicting depressive symptoms [10]. Validating models on harmonized datasets helps reduce overfitting and enhances clinical utility.

Longitudinal data plays a central role in osteoporosis prediction because it captures changes in BMD and related factors (e.g., age, weight, physical activity) over time, improving risk estimation [11]. It also reveals early bone loss, gradual declines in bone mass, and the influence of gender and lifestyle behaviors on BMD [12]. Incorporating these dynamic features enables ML models to produce more accurate and clinically relevant predictions.

Gender differences are key in osteoporosis modeling. Women experience faster bone loss, especially after menopause, while men exhibit slower declines [13]. Longitudinal data captures these differences and improves risk assessment [14]. However, many studies still combine male and female data, limiting model performance [15]. Incorporating clinical factors such as age, weight, and fracture history improves ML-based prediction, with longitudinal models showing the greatest benefit [16].

Diabetes and lifestyle factors also influence bone health. Although type 2 diabetes is often associated with higher BMD, it can reduce bone turnover and increase fragility [17]. Behaviors such as physical activity, diet, smoking, and alcohol use shape BMD trajectories over time. Resistance training, for example, improves BMD in older adults [18]. Despite these known relationships, many ML models omit diabetes and lifestyle features, limiting predictive strength.

This study uses dynamic, longitudinal data to create separate ML models for men and women, recognizing that differences in bone structure, metabolism, and how osteoporosis progresses can vary significantly by gender. The goal is to build predictive models that can identify the risk of osteoporosis and fractures early, paving the way for personalized care. By focusing on long-term changes and key risk factors like Physical functions and medical data, the study aims to improve how we assess and manage osteoporosis, reduce the likelihood of fractures, and ultimately enhance patient well-being while easing the strain on healthcare systems.

## 2. Methods

### 2.1 Dataset Description

We worked with two datasets, one for females and one for males, with data collected over several visits. Female Dataset: The SOF [7] tracked women 65 and older in four American cities between 1986 and 2017. With an emphasis on osteoporosis risk across time, the dataset comprises approximately 10,000 participants and collects data on BMD, physical examinations, and questionnaires every four months. Even though the SOF study calculated T-scores at Visit 2 and beyond using patients’ femoral neck BMD values to determine osteoporosis status, these T-scores were not directly recorded in the dataset. Male Dataset: The MrOS study [8], which started in 1998 with 5,994 enrollments from the US, concentrated on males 65 and older. More participants were added later, and the project was extended globally to examine older men’s health and bone related data over time. In order to evaluate osteoporosis and related diseases, it comprises baseline measurements of BMD and medical history, followed up with data every four months, and thorough exams every five years. In contrast to SOF study, the MrOS study included T-scores since Visit 1 in the database to explicitly identify patients’ osteoporotic status.

#### Data Pre-processing

As shown in Figure 1, two visits were selected for male and female participants by choosing the pairs with the highest overlap in patient IDs and shared features. This ensured consistency across visits and reduced bias from mismatched or incomplete data. Consequently, Visit 2 (V2) and Visit 5 (V5) were used for the female dataset, while Visit 1 (V1) and Visit 2 (V2) were selected for the male dataset.

**Figure 1:**
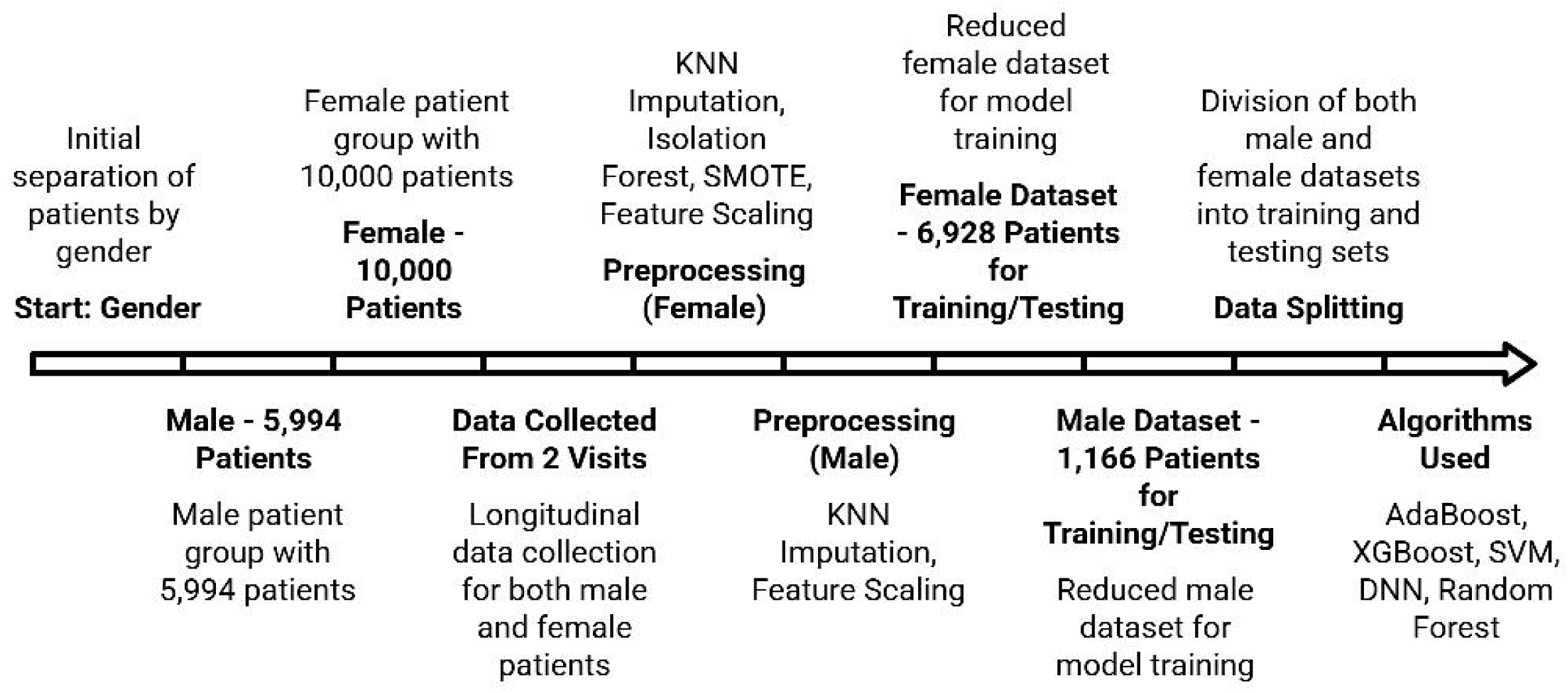
Osteoporosis prediction workflow.

Data preprocessing included handling missing values, updating column labels, detecting outliers, and addressing class imbalance. Missing values were imputed using KNN (K=5), which predicts values based on the five nearest neighbors and captures local data patterns more effectively than mean or mode imputation [19]. Column names were appended with visit identifiers (e.g., “V1”), and datasets were merged using participant IDs, retaining only individuals with complete data across both visits.

After feature engineering, outliers were removed using the Isolation Forest algorithm with a 20% contamination rate [20], reducing the influence of extreme values and improving data robustness. To address class imbalance, the Synthetic Minority Over-sampling Technique (SMOTE) was applied to generate synthetic minority samples and improve class distribution. These preprocessing steps ensured that the final dataset was clean, consistent, and ready for analysis.

### 2.2 Feature Engineering

#### 2.2.1 Feature Selection

A set of features with relevance to osteoporosis risk was selected, focusing on clinical, demographic, and lifestyle factors for both male and female participants as shown in the Table 1. These features were chosen to ensure a comprehensive assessment of bone health over different visits.

**Table 1:** Feature Example for Male and Female Participants.

| **Feature** | **Gender** |
| --- | --- |
| Height | both |
| BMD | both |
| Diabetes | both |
| Femoral Neck BMD T-score | male |
| Fasting Glucose | male |
| Testosterone | male |
| Weight | female |
| Menopause | female |
| Vertebral Fracture | female |

#### 2.2.2 Delta Variables for Longitudinal Analysis

To capture changes over time, we introduced new columns representing delta values, calculated as the difference between feature values at two time points. These delta variables reflect the progression or regression of key features, providing valuable insights into the temporal dynamics of osteoporosis risk factors. The delta values were computed as:

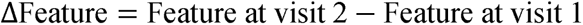

By including these delta variables, we can examine how changes in various factors such as weight, BMD, and lifestyle habits over time influence the likelihood of developing osteoporosis. This approach provides a dynamic view of health, helping to reveal trends and patterns that may not be visible from the static feature values alone. This method gives us important insights into how osteoporosis progresses over time and helps us better understand how changes in these parameters affect the chance of developing the condition.

#### 2.3.3 Binary Feature Consolidation

For binary features, the final value was set to 1 if the value was 1 at either visit. This approach ensures that any occurrence of a condition or behavior (e.g., smoking, fracture history) is captured, reflecting its clinical relevance to osteoporosis risk assessment.

### 2.3 Statistical Analysis using T-Test

A statistical analysis was conducted to identify the most significant features associated with hip osteoporosis in male and female participants prior to model development. For each dataset, Pearson correlation coefficients were computed between numeric features and the target variable to examine linear associations [21]. Independent two-sample t-tests (Welch’s correction for unequal variances) [22] were then performed to compare feature means between osteoporosis and non-osteoporosis groups. The resulting t-statistics, p-values, and correlation coefficients were combined into a single summary table, (Table 2) for each gender.

**Table 2:** Top Statistical Features by Gender with T-Test and Correlation Results. Note: Feature names in the table are coded. See Appendix B for the description mapping of the feature names.

| **Gender** | **Feature** | **T-Statistic** | **P-Value** | **Correlation** |
| --- | --- | --- | --- | --- |
| Male | v1_TBD | 5.429 | < 0.001 | -0.1359 |
|  | v1_L1D | 9.838 | < 0.001 | -0.2365 |
|  | v1_L2A | 3.959 | < 0.001 | -0.1145 |
|  | v1_L2C | 9.625 | < 0.001 | -0.2340 |
|  | v1_L2D | 10.526 | < 0.001 | -0.2526 |
|  | v1_L3C | 7.107 | < 0.001 | -0.1882 |
|  | v1_L3D | 8.412 | < 0.001 | -0.2121 |
|  | v1_L4C | 7.152 | < 0.001 | -0.1853 |
|  | v1_L4D | 8.491 | < 0.001 | -0.2146 |
|  | v1_HDC | 4.751 | < 0.001 | -0.1283 |
| Female | v2_WGHT | 4.143 | < 0.001 | -0.0493 |
|  | v5_HHR1 | -8.978 | < 0.001 | 0.1131 |
|  | v5_CLBR1 | -7.545 | < 0.001 | 0.1026 |
|  | v5_WLKR1 | -8.892 | < 0.001 | 0.1188 |
|  | v5_HIPTOV | -4.420 | < 0.001 | 0.0669 |
|  | v5_ANYTOV2 | -10.978 | < 0.001 | 0.1254 |
|  | v5_OSTFX | -15.685 | < 0.001 | 0.2565 |
|  | v5_VERT | -4.255 | < 0.001 | 0.0708 |
|  | v5_DOCF | -5.750 | < 0.001 | 0.0827 |
|  | v5_HIPXF | 5.179 | < 0.001 | -0.0620 |

Given the large number of variables (155 for females and 142 for males), only the top ten features with the most statistically significant differences (smallest p-values) were reported to highlight the most relevant associations.

### 2.4 Data Splitting

The datasets associated with male and female participants were split 80-20, with 80 percent of the data reserved for training and 20 percent reserved for testing as shown in the Table 3. This approach helped maintain an unbiased evaluation of the models’ predictive performance. Stratification was applied where necessary to ensure the distribution of key factors like gender, age, and comorbidities (e.g., diabetes) remained balanced.

**Table 3:**
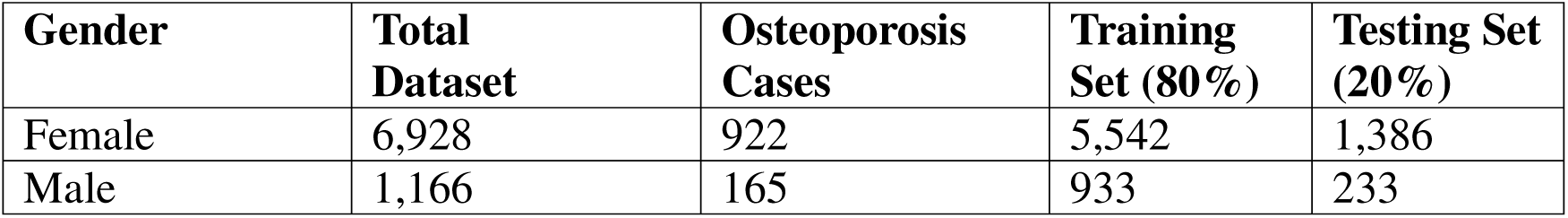
Dataset Splitting for Model Training and Testing.

### 2.5 Algorithms

The study employed five ML algorithms like eXtreme Gradient Boosting (XGBoost), Support Vector Machine (SVM), Adaptive Boosting (AdaBoost), Random Forest (RF), and Deep Neural Networks (DNN) to predict osteoporosis risk (Figure 1). These models were selected to capture longitudinal patterns and gender-specific variations using diverse computational approaches.

Each algorithm was evaluated for its ability to model non-linear relationships, temporal changes, and gender specific feature interactions. As detailed in the results, XGBoost performed particularly well for females, while RF captured male-specific patterns, highlighting the value of choosing algorithms suited to the characteristics of each dataset.

**XGBoost** [23] uses gradient boosting to capture complex interactions, making it well suited for the high-dimensional longitudinal features in this study. It demonstrated strong performance, particularly for the female dataset, aligning with previous applications in health research [24]. **SVM** separates classes using optimal hyperplanes and kernel functions [25], which helped model non-linear patterns in the male dataset.

**AdaBoost** iteratively emphasizes misclassified instances [26], allowing it to detect subtle clinical differences relevant to osteoporosis risk.

**DNNs** learn hierarchical representations [27], enabling them to capture complex temporal patterns, though they required careful regularization due to dataset size constraints.

**RF** combines multiple decision trees [28] and performed strongly for male participants, effectively modeling heterogeneous clinical and DXA related features.

### 2.7 Hyperparameter Tuning with Bayesian optimization

Bayesian optimization was used for hyperparameter tuning, requiring fewer evaluations than grid search [29]. Thirty-five iterations were run, five random and thirty guided, improving accuracy, F1 score, and Area Under the Curve (AUC) - Receiver Operating Characteristic (ROC). Gender-specific tuning further enhanced model performance. Hyperparameter bounds are summarized in Table 4.

**Table 4:**
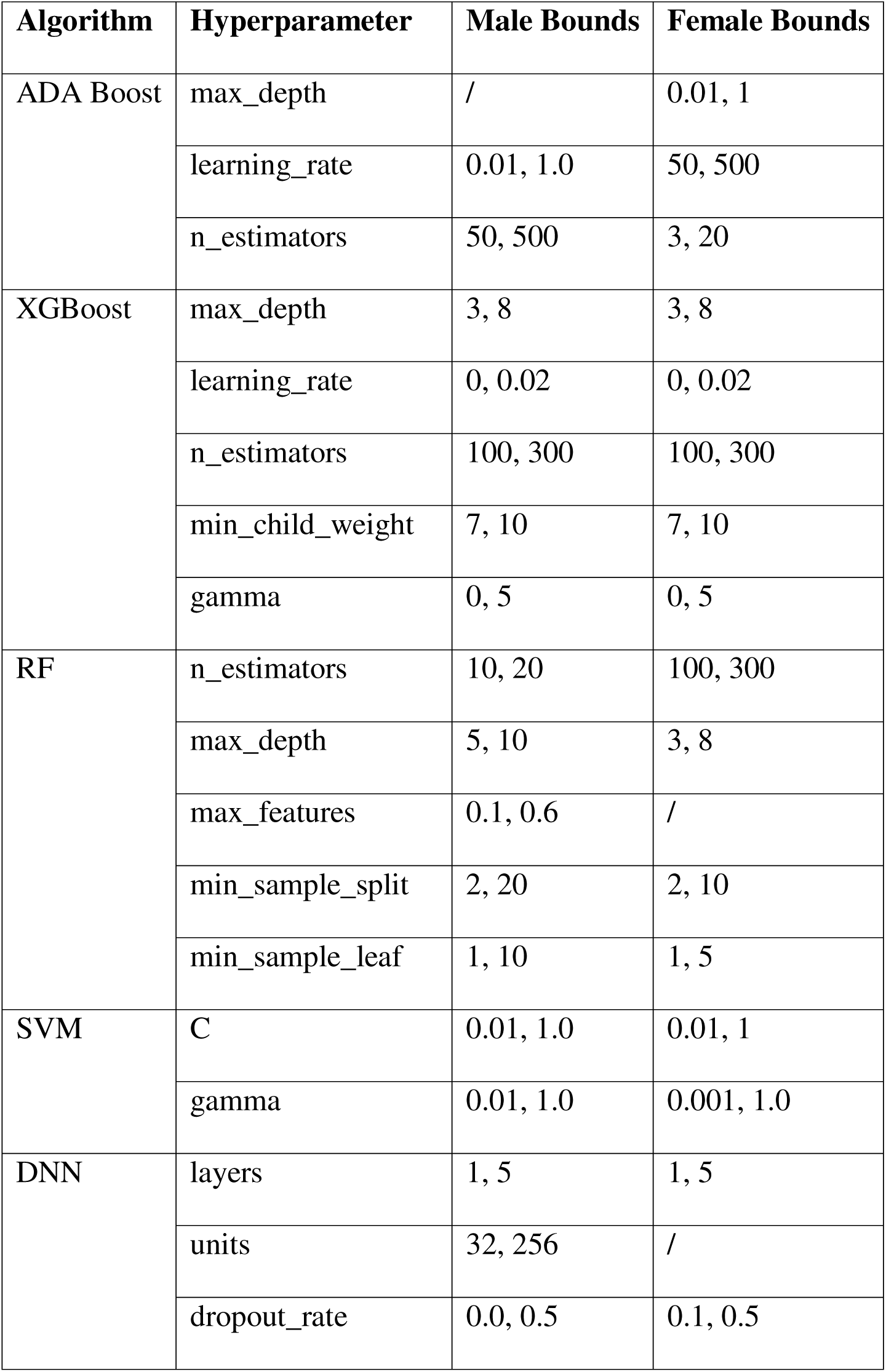
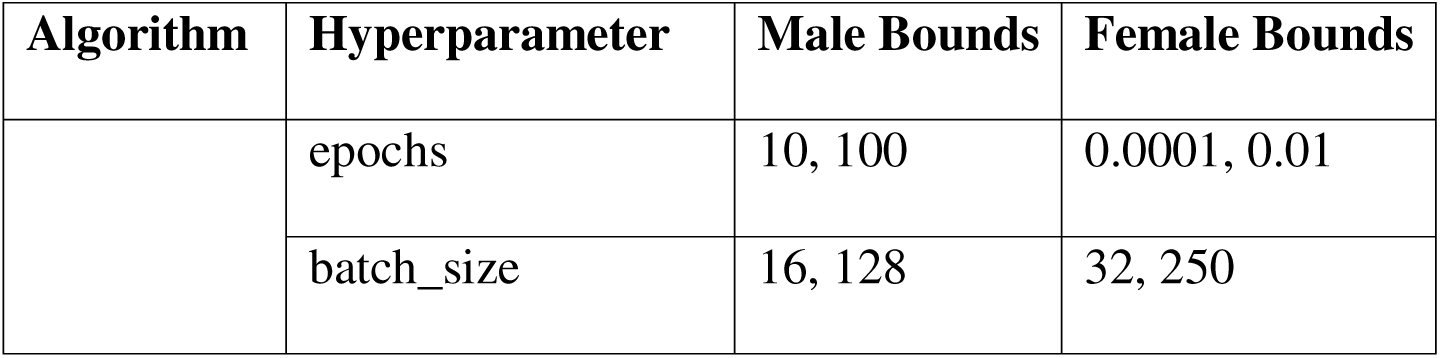
Hyperparameters and Bounds for Algorithms.

| **Algorithm** | **Hyperparameter** | **Male Bounds** | **Female Bounds** |
| --- | --- | --- | --- |
| ADA Boost | max_depth | / | 0.01, 1 |
|  | learning_rate | 0.01, 1.0 | 50, 500 |
|  | n_estimators | 50, 500 | 3, 20 |
| XGBoost | max_depth | 3, 8 | 3, 8 |
|  | learning_rate | 0, 0.02 | 0, 0.02 |
|  | n_estimators | 100, 300 | 100, 300 |
|  | min_child_weight | 7, 10 | 7, 10 |
|  | gamma | 0, 5 | 0, 5 |
| RF | n_estimators | 10, 20 | 100, 300 |
|  | max_depth | 5, 10 | 3, 8 |
|  | max_features | 0.1, 0.6 | / |
|  | min_sample_split | 2, 20 | 2, 10 |
|  | min_sample_leaf | 1, 10 | 1, 5 |
| SVM | C | 0.01, 1.0 | 0.01, 1 |
|  | gamma | 0.01, 1.0 | 0.001, 1.0 |
| DNN | layers | 1, 5 | 1, 5 |
|  | units | 32, 256 | / |
|  | dropout_rate | 0.0, 0.5 | 0.1, 0.5 |

| Algorithm | Hyperparameter | Male Bounds | Female Bounds |
| --- | --- | --- | --- |
|  | epochs | 10, 100 | 0.0001, 0.01 |
|  | batch_size | 16, 128 | 32, 250 |

## 3 Results

### 3.1 Dataset Overview

The female dataset, after preprocessing, includes 6,928 individuals, with 922 diagnosed with osteoporosis, as shown in Table 3. For each visit, 51 features were extracted, and 50 delta features were added to capture changes over time. A column was also included to indicate whether the individual had osteoporosis.

The male dataset, after preprocessing, consists of 1,166 individuals, with 165 diagnosed with osteoporosis. From each visit, 55 features were extracted, and 38 delta features were created. A column indicating osteoporosis status was also included.

### 3.2 Results of T-Test and Correlation Analysis

Table 2 presents the ten most statistically significant features for each gender. Among males, several lumbar spine BMD measures (v1_L2D, v1_L3D, v1_L4D) showed the strongest associations with hip osteoporosis, exhibiting high t-statistics (e.g., t = 10.526 for v1_L2D) and negative correlations, indicating that lower BMD was linked to a higher risk of osteoporosis.

In females, variables such as v5_OSTFX (osteoporotic fracture), v5_ANYTOV2 (any type of fracture), and v5_WLKR1 (walking ability) were most significant (all p< 0.001). Positive correlations for these features suggest that previous fractures and reduced mobility were associated with an increased risk of osteoporosis.

These results emphasize distinct gender specific patterns: BMD measures were dominant predictors in males, whereas fracture and mobility indicators were more influential in females.

### 3.3 Feature Importance in Predicting Osteoporosis

The top 20 features that are the main influences in XGBoost predictions for the female dataset are shown in Figure 2A. These features were selected for their importance in predicting osteoporosis risk. Physical functions, such as difficulty with heavy housework, walking, climbing stairs appeared as key factors. These activities are mostly linked to declining bone health, reduced mobility, and muscle strength, all of which contribute to osteoporosis development. The frequent occurrence of daily activity limitations underscores the importance of functional impairment in identifying at-risk individuals.

**Figure 2:**
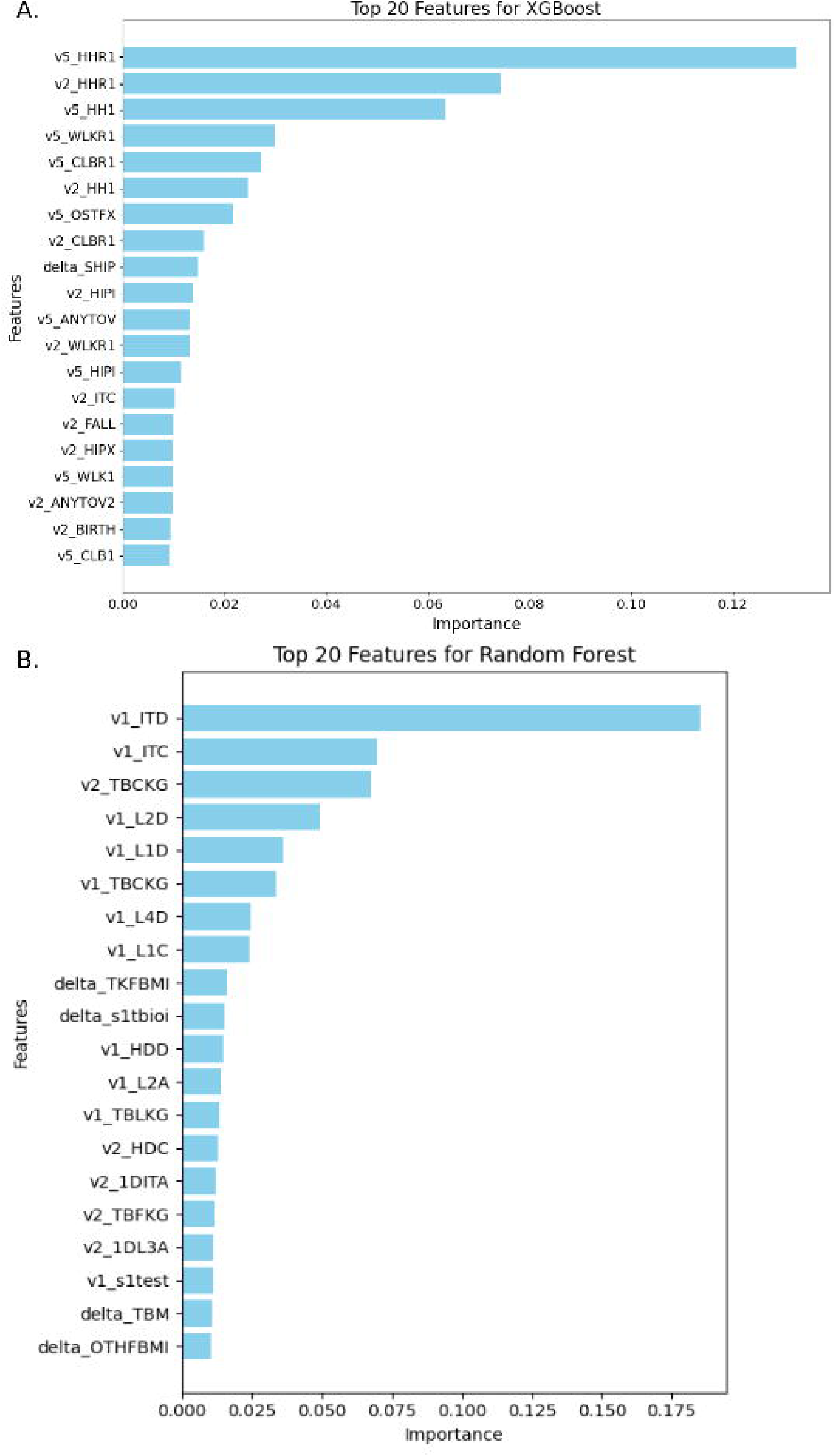
(A) Top 20 features influencing XGBoost predictions for osteoporosis risk in female (B) Top 20 features influencing RF predictions for osteoporosis risk in male. Note. Feature names in the figure are coded. See Appendix B for the description mapping of the feature names.

Additionally, fracture history, especially vertebral and hip fractures, was a strong predictor of osteoporosis. Previous fractures indicate weakened BMD, increasing the likelihood of future fractures. Features related to changes in hip pain and overall health further highlight the role of dynamic health changes in osteoporosis progression. These findings emphasize the critical role of both functional limitations and fracture history in understanding and predicting osteoporosis risk.

Figure 2B shows a visualization of the top 20 features in order of importance to the male RF model. Physical features are shown to have the greatest influence, with corrected Intertroch BMD taking strong precedence followed by Intertrochanteric BMC and Total Body BMC.

It is clear that features related to BMD and BMC are the most prominent indicators of osteoporosis risk in male participants. The figure further demonstrates that features indicating changes in testosterone levels are also shown to have a notable impact on male osteoporosis risk.

### 3.4 Model Performance by Algorithm

The performance of various ML algorithms is summarized in Table 5. For the female dataset, all algorithms showed strong performance, with XGBoost achieving the highest F1 score of 0.874. SVM and DNN also performed well, with F1 scores of 0.820 and 0.872, respectively.

**Table 5:** Performance Metrics for Different Algorithms by Gender.

| Algorithm | Gender | F1 Score | Recall | Precision | Accuracy (%) | AUC-ROC |
| --- | --- | --- | --- | --- | --- | --- |
| AdaBoost | Female | 0.854157 | 0.854891 | 0.859307 | 85.00 | 0.917561 |
| AdaBoost | Male | 0.799055 | 0.862661 | 0.744184 | 86.00 | 0.802550 |
| XGBoost | Female | 0.874472 | 0.875000 | 0.878781 | 88.00 | 0.938857 |
| XGBoost | Male | 0.808589 | 0.854077 | 0.794433 | 85.00 | 0.836521 |
| RF | Female | 0.824536 | 0.825000 | 0.826622 | 82.00 | 0.902707 |
| RF | Male | 0.860462 | 0.846154 | 0.888419 | 85.00 | 0.894802 |
| SVM | Female | 0.820000 | 0.800000 | 0.840000 | 81.00 | 0.910000 |
| SVM | Male | 0.791209 | 0.857143 | 0.734694 | 86.00 | 0.500000 |
| DNN | Female | 0.872504 | 0.872826 | 0.874732 | 87.28 | 0.929723 |
| DNN | Male | 0.799222 | 0.862779 | 0.744387 | 86.00 | 0.500000 |

For the male dataset, RF provided the best overall performance with scores that stood out in all categories, culminating in an F1 score of 0.860462. XGBoost showed the second-best performance, though the metrics noticeably differed from RF in distribution. The remaining algorithms, AdaBoost, SVM, and DNN were somewhat lacking in performance with lower precision and F1 scores despite having decent recall.

### 3.5 Accuracy and AUC-ROC Metrics

The performance metrics of various algorithms for predicting osteoporosis risk, based on Accuracy and AUC-ROC, are shown in Table 4. The algorithms performed well on the female dataset, with XGBoost achieving the highest accuracy (88%) and AUC-ROC (0.9389), followed by AdaBoost with an accuracy of 85% and AUC-ROC of 0.9176. Other algorithms, such as DNN and SVM, also performed strongly, with DNN achieving an accuracy of 87.28% and an AUC-ROC of 0.9297, and SVM reaching an accuracy of 81% and AUC-ROC of 0.9100. The ROC curve in Figure 3A displays the performance of the XGBoost model specifically for the female dataset. The curve shows a high rate of true positives and a low rate of false positives, indicating that the model is highly proficient at differentiating between osteoporosis and non-osteoporosis cases. With an AUC of 0.93, the model’s performance is strong, indicating a reliable ability to correctly classify individuals with and without osteoporosis.

**Figure 3:**
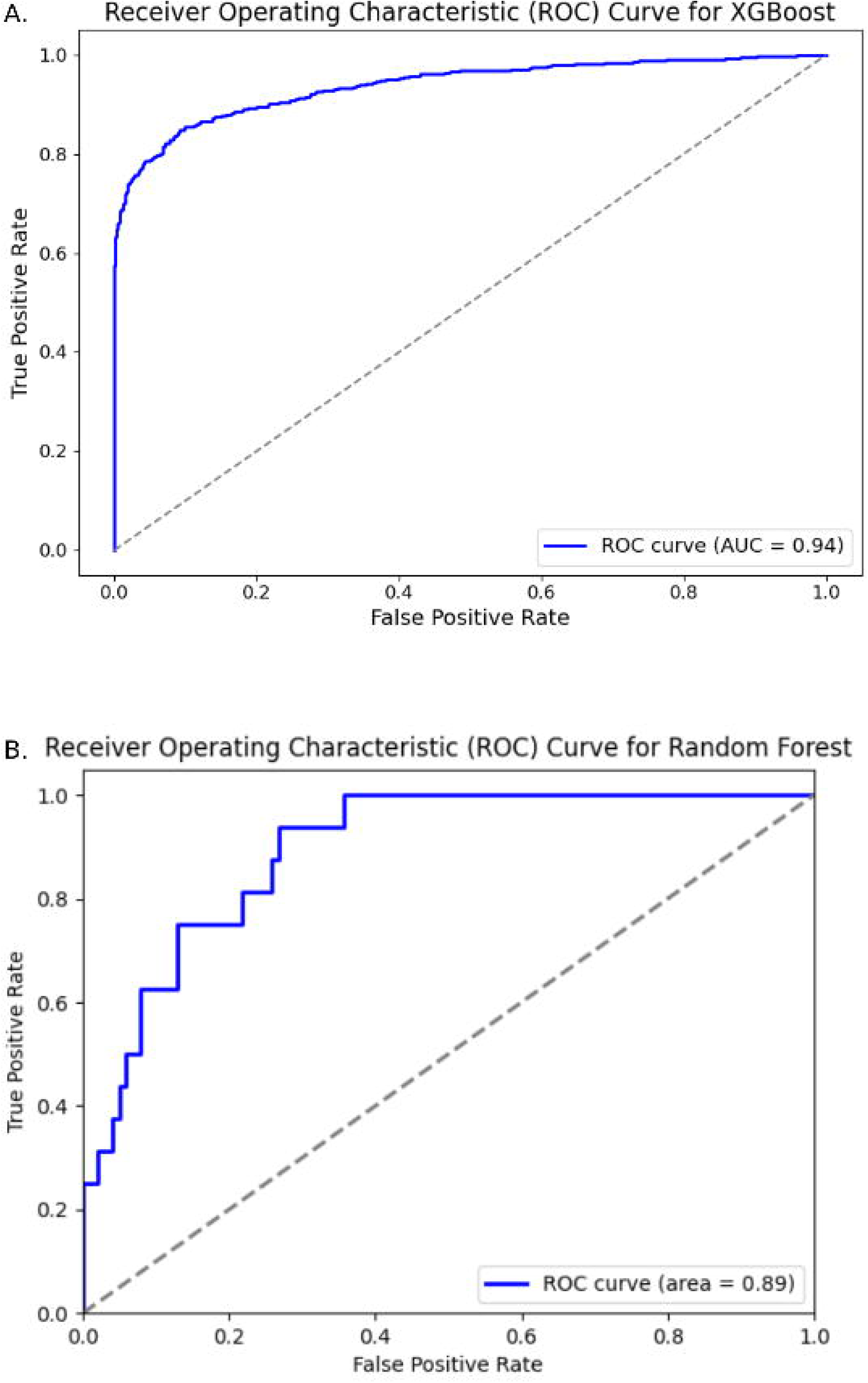
(A) ROC Curve for XGBoost Model Performance for Female dataset (B) ROC Curve for RF Model Performance for Male dataset.

For the male dataset, AdaBoost, SVM, and DNN showed similar accuracy of 86%, though the AUC-ROC scores of SVM and DNN suggest that they found it especially challenging to distinguish between examples within the imbalanced male dataset.

For this reason, it can be concluded that they were not as effective in predicting osteoporosis risk for male participants. With only a small decrease in accuracy at 85%, RF stood out significantly with an AUC-ROC score of 0.8948, which is visualized in Figure 3B. As a result, RF serves as the best combination of accuracy and AUC-ROC. Overall, the metrics reveal that the male dataset was more of a challenge for the models, possibly due to the lesser number of positive cases and records as a whole.

### 3.6 Confusion Matrix

The confusion matrix for the model’s performance with the female and male dataset are shown in Figure 4A. For Female data, XGboost confusion matrix showed a higher number of true positives and relatively less false positives, which indicates the accurate identification of individuals with osteoporosis. While there are some false negatives, suggesting a potential area for improvement in detecting all true cases, the overall performance remains strong.

**Figure 4:**
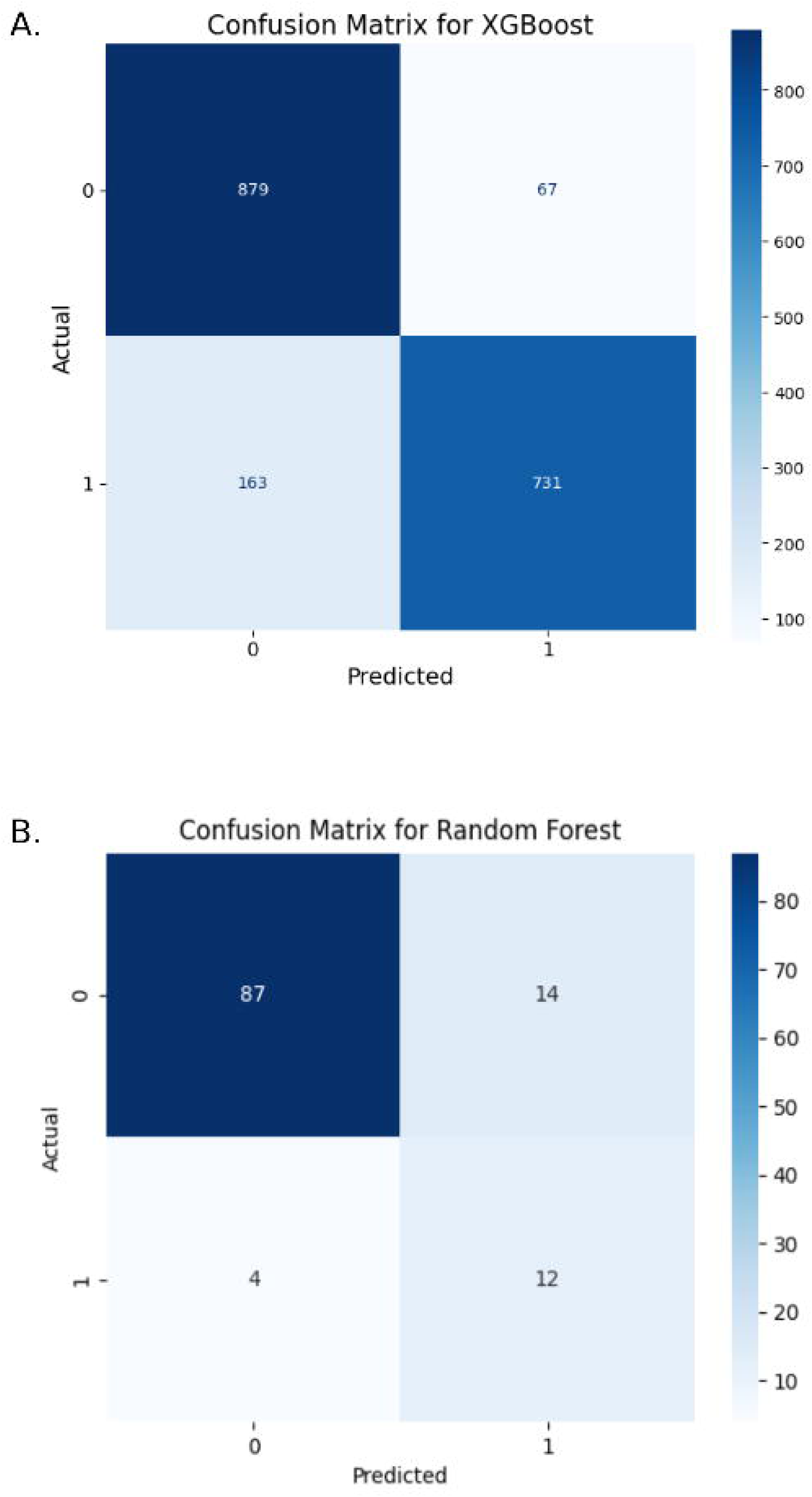
(A) Confusion Matrix for XGBoost (Female dataset) (B) Confusion Matrix for RF (Male dataset).

The confusion matrix for the male RF model is shown in Figure 4B. This model showed greater accuracy with smaller test sets, hence the lower number of examples shown in the matrix. Figure 4B demonstrates that the model was generally less effective in classifying positive cases, which is to be expected as there were fewer examples of such.

## 4 Discussion

This study demonstrates the value of ML for predicting osteoporosis using multi-visit clinical data. Longitudinal features, especially delta variables improved predictive accuracy by capturing temporal trends. Ensemble and deep learning models, particularly XGBoost and RF, effectively modeled nonlinear and gender-specific relationships.

Limitations include demographic imbalance, missing clinical data requiring imputation, and the absence of genetic and biochemical markers. Future work should incorporate additional visits, biomarkers, and medication histories, and validate models across diverse populations. Collaboration with clinicians will support the development of practical tools for real-world use.

In this study, several machine learning models were evaluated for use in predicting the risk of osteoporosis, introducing the use of longitudinal data in a gender-specific approach. These experiments provided valuable insights into the nuances of ML model training as well as the impact of gender-specific features in evaluation of osteoporosis risk. For the female dataset, the XGBoost model achieved strong performance with high accuracy and AUC-ROC scores, making it an effective tool for predicting osteoporosis risk in women by incorporating time-based health changes. For the male dataset, the RF outperformed the others, effectively capturing patterns in longitudinal data specific to men.

In conclusion, with consideration for changes in bone health, physical functions, and medical history over time, these gender-specific models improve prediction accuracy and underscore the importance of acknowledging gender-related differences in osteoporosis risk assessment. This approach shows promising potential for the improvement of early detection and personalized management of osteoporosis, with further optimization required to improve model performance between genders.

## Author contributions

**Shyama Prasad Tripathy:**Writing – original draft, Methodology, Formal analysis, Data curation, Validation. **LohithaSaripalli:**Writing – original draft, Formal analysis, Data curation. **Katherine Berry:**Writing – original draft, Formal analysis, Data curation. **A. Champa Jayasuriya:** Conceptualization, Supervision, Investigation,Writing – review and editing. **Devinder Kaur:** Supervision, Investigation, Resources. **Fayeq Jeelani Syed:** Conceptualization, Supervision,Investigation, Writing – review and editing.

## Ethics Statement

This study used SOF and MrOS databases and did not involve any human participants, or patient data.

## Data availability

The datasets and codes available at https://github.com/Sprasad2058/Enhancing-Osteoporosis-Risk-Prediction-Gender-Based-Models-from-Longitudinal-Data

## Appendix A: Variable Description Mapping of Important Features

### Appendix A.1. Female Top 20 Important Features

v5_HHR1 = V5 Do you have difficulty doing heavy housework?

v2_HHR1 = V2 Do you have difficulty doing heavy housework?

v5_HH1 = V5 Can you do heavy housework?

v5_WLKR1 = V5 Difficulty in walking

v5_CLBR1 = V5 Difficulty in climbing step

v2_HH1 = V2 Can you do heavy housework?

v5_OSTFX = V5 Vertebral fracture

v2_CLBR1 = V2 Difficulty in climbing step

delta_SHIP = delta Hip pain

v2_HIPI = V2 Hip fracture post current visit

v5_ANYTOV = V5 Any fracture since age 50

v2_WLKR1 = V2 Difficulty in walking

v5_HIPI = V5 Hip fracture post current visit

v2_ITC = V2 Intertrochanteric BMC

v2_FALL = V2 Fallen or hit an object

v2_HIPX = V2 Hip fracture post current visit

v5_WLK1 = V5 Can you walk 2-3 blocks outside?

v2_ANYTOV2 = V2 Any fracture since age 50

v2_BIRTH = V2 Have you ever given birth?

v5_CLB1 = V2 Can you climb 10 steps without stopping?

### Appendix A.2. Male Top 20 Important Features

V1_ITD = V1 Corrected Intertroch BMD (g/cm^2^)

V1_ITC = V1 Intertrochanteric BMC (g)

V2_TBCKG = V2 Total body BMC (kg)

V1_L2D = V1 L2 BMD (g/cm^2^)

V1_L1D = V1 L1 BMD (g/cm^2^)

V1_TBCKG = V1 Total body BMC (kg)

V1_L4D = V1 L1 BMD (g/cm^2^)

V1_L1C = V1 L1 BMC (g)

Delta_TKFBMI = difference in Trunk fat BMI (kg/m^2^)

Delta_s1tbioi = difference in Bioavailable T- nmol/l

V1_HDD = V1 head BMD (g/cm^2^)

V1_L2A = V1 L2 Area (cm^2^)

V1_TBLKG = V1 Total body lean mass (kg)

V2_HDC = V2 head BMC (g)

V2_1DITA = V2 Intertrochanter Area abs change since V1

V2_TBFKG = V2 Total body fat mass (kg)

V2_1DL3A = V2 L3 Area (cm2) abs change since V1

V1_s1test = V1 Testosterone (ng/dl)

Delta_TBM = difference in Total Mass (g)

Delta_OTHFBMI = difference in Non-Trunk fat BMI (kg/m^2^)

## Appendix B. Variable Description Mapping of Statistical Features

### Appendix B.1. Male top Statistical Features with T-Test and Correlation Results

v1_TBD = Whole Body Total BMD (g/cm^2^)

v1_L1D = V1 L1 BMD (g/cm^2^)

v1_L2A = V1 L2 Area (cm^2^)

v1_L2C = V1 L2 BMC (g)

v1_L2D = V1 L2 BMD (g/cm^2^)

v1_L3C = V1 L3 BMC (g)

v1_L3D = V1 L3 BMD (g/cm^2^)

v1_LAC = V1 Left Arm BMC (g)

v1_LAD = V1 Left Arm BMD (g/cm^2^)

v1_HDC = Head BMC (g)

### Appendix B.2 Female top Statistical Features with T-Test and Correlation Results

v2_WGHT = V2 Current weight (kg)

v5_HHR1 = V5 Recoded used for summary1: Do you have difficulty doing heavy house work? Y/N

v5_CLBR1 = V5 Recoded used for summary1: Do you have difficulty climbing up 10 steps without stopping/resting? Y/N

v5_WLKR1 = V5 Recoded used for summary1: Do you have difficulty walking 2-3 blocks outside on level ground? Y/N

v5_HIPTOV = V5 Adjudicated: hip fracture since age 50?

v5_ANYTOV2 =V5 Adjudicated: any fracture since age 50?

v5_OSTFX = V5 Osteoporosis or vertebral fracture (self report)

v5_VERT=V5 Has a doctor (or health care provider) ever (V1) (or since you last visited our clinic (about 2 years ago) (V2), or during the past 12 months (A3), or since you last completed a questionnaire for the study (V3, V4, V8), or in the last 2 years (V5, V6), or since your last visit (V7) told you that you had a fracture of the spine or fracture of the vertebrae? (self report)

v5_DOCF = V5 Has your doctor (or health care provider) ever (V1, AA) (or, since you last completed a questionnaire for our study (about 12 months ago) (V2, V3, V4), or in the last two years (V5, V6), or since your last visit (V8, V9)) said that you had a broken or fractured bone? (self report)

v5_HIPFX = V5 Follow-up time (in days) to 1st hip fracture, excluding prior hip fracture, since current visit

